# Epidemiology, Temporal and Seasonal Trends, and Geographic Distribution of Leptospirosis in the Dominican Republic, 2012–2026

**DOI:** 10.64898/2026.09.02.26362038

**Authors:** Lisette V. Alcántara, Jose J. Sánchez, David De Luna, O. Alejandro Aleuy, Benjamin L. Miller, Charles R. Mace, Hunter A. Scott, Lara Smejkal, José L. Cruz, Charles H. Hennekens, Timothy D. Dye

## Abstract

Leptospirosis is a widespread zoonotic infection and a growing public health concern in tropical, resource-limited settings, yet remains underrecognized and underreported where diagnostic and surveillance infrastructure is limited. We conducted an analytical cross-sectional study of national surveillance data from the Dominican Republic (2012–June 2026), integrating geospatial meteorological data to characterize disease burden, trends, seasonality, and clinical risk factors. Of 8,425 records, 5,412 met case-definition criteria (3,441 suspected, 1,448 probable, 523 confirmed), yielding a cumulative incidence of 3.55 per 100,000 person-years (2012–2025). Laboratory confirmation rose markedly, from 9.7% overall to half of 2025 cases and over half in 2026. Overall incidence remained stable across all years (p = 0.15), though with three distinct periods: declining infection pre-COVID, low infection during COVID, and increasing infection post-COVID. Cases clustered in the rainy/hurricane season (61.1%, May–November; p< 0.001), with a marginal rainfall correlation (r= 0.553, p= 0.062). Rural provinces bore disproportionate risk, led by Hermanas Mirabal (16.43/100,000 person-years), while large urban and coastal provinces had rates 80% lower. Men had 2.73-fold higher incidence than women (95% CI 2.56– 2.90), peaking at younger ages 10–29. Comorbidity was the strongest predictor of complications (OR 2.84, 95% CI 2.01–4.02, p< 0.001). Limited diagnostic confirmation remains the central obstacle to characterizing leptospirosis burden in the Dominican Republic, with confirmation rates exceeding 40% only from 2023 onward. This first multi-decade national analysis identifies high-risk provinces, demographic groups, and seasonal windows for targeted surveillance, highlighting expanded diagnostic capacity as a priority for burden estimation and case management.

## Introduction

Leptospirosis is one of the most widespread zoonotic infections worldwide and represents an important public health concern, particularly in tropical and resource-limited settings.^1,2,3^ Leptospirosis is caused by *Leptospira spp*., and infection ranges from a self-limited febrile illness to severe disease, including Weil’s disease, pulmonary hemorrhage, multiorgan failure, and death.^4,5,6^ The estimated global burden is approximately 1.03 million cases and 58,900 deaths annually, corresponding to roughly 2.90 million disability-adjusted life years (DALYs) lost. Leptospirosis remains underrecognized and underreported, particularly where access to confirmatory diagnostics and surveillance infrastructure is limited.^7,8^ Hospital-based surveillance may capture only a fraction of infections, further complicating efforts to characterize disease patterns and identify the factors driving transmission.^2,9^

Transmission is closely tied to environmental and occupational exposure. Regional and global reviews consistently identify contact with animal reservoirs, freshwater, and soil as central to transmission,^3,10^ with risk shaped by water contact, sewage proximity, livestock proximity, sanitation access, and occupation in both rural and urban settings.^10,11^ Male gender, particularly among economically active adults, is consistently identified as an independent risk factor, likely reflecting greater exposure through occupations such as agriculture, animal husbandry, and construction, as well as recreational water contact.^10–12^ Similarly, a study in the Dominican Republic found higher seroprevalence among males than females (17.8% vs. 7.7%; adjusted OR, 2.41; 95% CI, 1.79– 3.25).^13^ Increasing age was also associated with higher seroprevalence, rising from 2.8% among participants aged 5–19 years to 13.7% among those aged 35–49 years and 19.3% among those aged ≥65 years.^13^ However, in other tropical settings, young adults may face greater risk due to increased occupational, school-related, recreational, and travel-related water/soil exposures during working and school years, with risk declining with age as environmental exposure decreases and acquired immunity accrues.^14,15^ Recreational water exposure, in particular, has been associated with outbreaks among younger populations, contrasting with the primarily occupational exposures observed among older adults.^16^ Whether the Dominican age distribution follows the older pattern reported in the national seroprevalence survey or the younger pattern described elsewhere in the tropics has not been established from surveillance data.

Climate and rainfall play well-documented roles in shaping transmission dynamics. Rainfall and flooding increase opportunities for exposure by facilitating the presence of contaminated surface water, disrupting sanitation, and resuspending *Leptospira* from soil, typically producing a lag of days to weeks between precipitation events and case peaks.^17,18,19,20,21,22,23,24^ Beyond seasonal rainfall, extreme events such as hurricanes and severe flooding can rapidly alter environmental conditions and disrupt infrastructure, creating conditions for localized outbreaks.^21,25,26,27,28^ Broader climate variability, including El Niño–Southern Oscillation cycles and sea surface temperature, has also been linked to shifts in outbreak probability across the Pacific and Latin America.^29,30,31^ However, the strength and direction of these climate–disease relationships vary considerably by geography, underscoring the need to interpret them within local environmental and social context rather than generalizing across settings.^32,33,34,35,36,37,38,39^

The Caribbean is particularly relevant to this relationship given its regular exposure to hurricanes, heavy rainfall, and flooding, yet leptospirosis remains substantially understudied across the region, and its potential for serving as a nexus of infection for the Americas. A regional review identified persistent surveillance gaps and limited empirical evidence linking rainfall, flooding, and post-hurricane conditions to disease incidence.^40,41^ These gaps are particularly important in the Caribbean, where recurrent flooding, tropical cyclones, and other climate-related hazards can create environmental conditions conducive to leptospirosis transmission.^40^ Moreover, international tourism represents a distinct exposure pathway, with leptospirosis among returned travelers commonly linked to freshwater recreation in Southeast Asia, the Caribbean, and Central and South America.^42^ Given the major tourism sector of the Dominican Republic, travel-associated exposure may be an underrecognized transmission route alongside occupational and environmental exposures.^42^

The Dominican Republic offers a useful setting for exploring these interrelationships. The country has a tropical climate with seasonal rainfall, recurrent exposure to hurricanes, and areas affected by flooding and inadequate drainage, alongside agricultural and livestock activities, periurban expansion, and variable access to water and sanitation.^43,44^ Prior work suggests substantial underestimation of the national burden: a multistage seroepidemiologic survey estimated approximately 33,000 Leptospira infections annually nationwide, compared with an average of only 226 reported cases per year through existing surveillance during 2013–2022.^13^ Subsequent spatial analyses have identified province-specific environmental and socioeconomic correlates of risk, including freshwater exposure, river density, rainfall, and rat exposure.^43,44^

Much of this underestimation reflects diagnostic limitations rather than a truly low incidence. Confirmatory testing including microscopic agglutination testing (MAT) and polymerase chain reaction (PCR) requires specialized equipment and trained personnel that are not consistently available in affected settings; hence, diagnosis is often based on clinical suspicion alone, and reported cases likely skew toward more severe presentations.^1,6,8^ This diagnostic gap has direct clinical consequences: early antibiotic treatment is most effective when initiated before laboratory confirmation is typically available, but diagnostic delays may prevent timely treatment. Experience from outbreaks, including the 2009 Philippines flood, underscores the value of early clinical recognition and prompt treatment.^6,28^

In this analysis, we describe the epidemiology of leptospirosis in the Dominican Republic from 2012 to the present, examining patterns by person, place, and time. We also explore the seasonal distribution of cases in relation to national mean rainfall. Together, these findings help quantify the national burden of leptospirosis and inform surveillance, preparedness, and prevention efforts in the Dominican Republic.

## Materials and Methods

### Ethical Considerations

The Florida Atlantic University Institutional Review Board reviewed the protocol for this study (“Leptospirosis in the Dominican Republic, 2000–2026: National Surveillance Trends,” protocol IRB2606182) and determined it was exempt from federal regulation under 45 CFR 46.104(4)(ii) on June 22, 2026, given its nature as a secondary analysis of previously collected, de-identified surveillance data. The Dominican Republic’s Ministry of Public Health and Social Assistance (MISPAS) provided the data used in this analysis in anonymized, aggregate form, and the research team had no access to names, identification documents, or any other information that could directly identify patients.

### Study Design, Data Source, and Case Definition

We conducted descriptive analyses of national surveillance data from the Dominican Republic’s Ministry of Public Health and Social Assistance (MISPAS), combined with precipitation data from the Dominican Institute of Meteorology (INDOMET). We obtained case-level data from the Epidemiological Surveillance Information System (SIP-0276FA51, Leptospirosis, 2010– 2026), which the General Directorate of Epidemiology (DIGEPI) maintains. This system serves as the country’s official registry for mandatory leptospirosis reporting.

We received a de-identified extract containing 8,425 individual records with sociodemographic, clinical, laboratory, and final case classification variables from January 2012 through June 2026. Because 2026 data cover only the first half of the year, we explicitly flagged this incomplete period wherever it appears in estimates or analyses. The analysis covered the entire national territory at two levels of geographic disaggregation: the country’s 31 provinces and the National District, and, alternatively, the nine health regions defined by the National Health Service (SNS): 0 Metropolitana, I Valdesia, II Cibao Norte, III Cibao Nordeste, IV Enriquillo, V Este, VI El Valle, VII Cibao Occidental, and VIII Cibao Central.

Each record included a final classification variable distinguishing five mutually exclusive categories: suspected (1), probable (2), confirmed (3), discarded, and pending classification. We defined a valid leptospirosis case as any record classified as suspected, probable, or confirmed, following the clinical, epidemiological, and laboratory criteria of the national surveillance protocol. We excluded 2,827 records that lacked a final classification, because we could not determine their case status. The resulting analytic cohort comprised 5,412 valid cases: 3,441 suspected (63.6 %), 1,448 probable (26.8 %), and 523 confirmed (9.7 %). We assigned each case to a calendar year and month based on the reported date of symptom onset rather than the date of notification to better reflect the timing of disease occurrence.

### Statistical Analyses

We calculated annual incidence rates by dividing the number of cases with symptom onset in a given calendar year by the mid-year population estimate for that year in the Dominican Republic, expressed per 100,000 population. For rates by province, region, gender, or age group, we summed the numerator (cases) and denominator (person-years) in each year of the study period (2012–2025), rather than averaging annual rates, a standard approach in descriptive epidemiology that avoids overweighting years with small denominators. We also aggregated provinces into eight geographic zones (e.g., semi-arid Northwest, Cibao Valley, Metropolitan Santo Domingo) based on a qualitative, narratively assigned classification of physical geography and calculated zone-level incidence rates per 100,000 person-years with exact Poisson 95% confidence intervals. This exploratory aggregation was intended to identify broad spatial patterns for hypothesis generation; confirming environmental drivers will require future analyses using quantitative covariates (e.g., irrigation extent, livestock density, land cover) linked directly to surveillance data. We excluded 2026 from period rates and trend models because it is incomplete, although we report it descriptively on its own. We estimated the male-to-female incidence rate ratio and its 95% confidence interval using the normal approximation to the log rate ratio, with standard error equal to the square root of the sum of the inverses of the case counts in each group, a standard method for comparing incidence rates based on Poisson-type counting processes.

### Temporal Trends

We assessed the trend in the annual number of cases using ordinary least squares linear regression of the natural logarithm of the annual count on calendar year, restricted to the complete years of the study period (2012–2025). From the slope of this regression, we derived the annual percent change (APC), calculated as APC = (e&slope−1) × 100.^45^ For years with zero cases in each subcategory, we applied a continuity correction (+0.5) before the logarithmic transformation. We also fit an interrupted time-series model to monthly case counts, specifying two interruption points—COVID onset (early 2020) and resurgence onset (early 2023)—to allow level and trend to shift across three periods (pre-COVID, during-COVID, resurgence), distinguishing abrupt discontinuities (suggesting surveillance disruption) from gradual epidemiological change.

### Seasonal Variation

We assessed the presence of a seasonal pattern using a chi-square goodness-of-fit test, comparing the observed distribution of cases by calendar month (summed across all study years) against the null hypothesis of a uniform distribution across the twelve months. We also compared the proportion of cases occurring during the rainy/hurricane season (May–November) with the proportion occurring during the dry season (December–April).

### Multivariable Regression Models

We fitted a multivariable logistic regression model to identify factors associated with the presence of clinical complications, including gender, age group (reference: 20–29 years), presence of comorbidities, and year of occurrence (centered on the median year of the study period) as covariates. Model coefficients were estimated using the iteratively reweighted least squares algorithm (IRLS, Newton–Raphson), with standard errors derived from the Fisher information matrix. Results are expressed as odds ratios with 95% confidence intervals and Wald test p-values. We set statistical significance at a two-tailed p < 0.05 for all tests. We performed all data processing, structuring, and analyses, including exploratory analysis, incidence rate calculations, trend and seasonality tests, and logistic regression modeling, using IBM SPSS Statistics (version 29). We fit the interrupted time-series model in Python (version 3.14.7), and reported point estimates, 95% confidence intervals, and p-values as appropriate across all analyses.

## Results

### Cohort characteristics

Of the 8,425 records reported to the Epidemiological Surveillance Information System between 2012 and June 2026, we excluded 186 (2.2 %) that corresponded to discarded events and 2,827 (33.6 %) that were pending classification or missing this variable, leaving an analytic cohort of 5,412 valid cases. The cumulative incidence rate was 3.55 cases per 100,000 person-years during the study period (2012–2025) (Table 1).

**Table 1.**
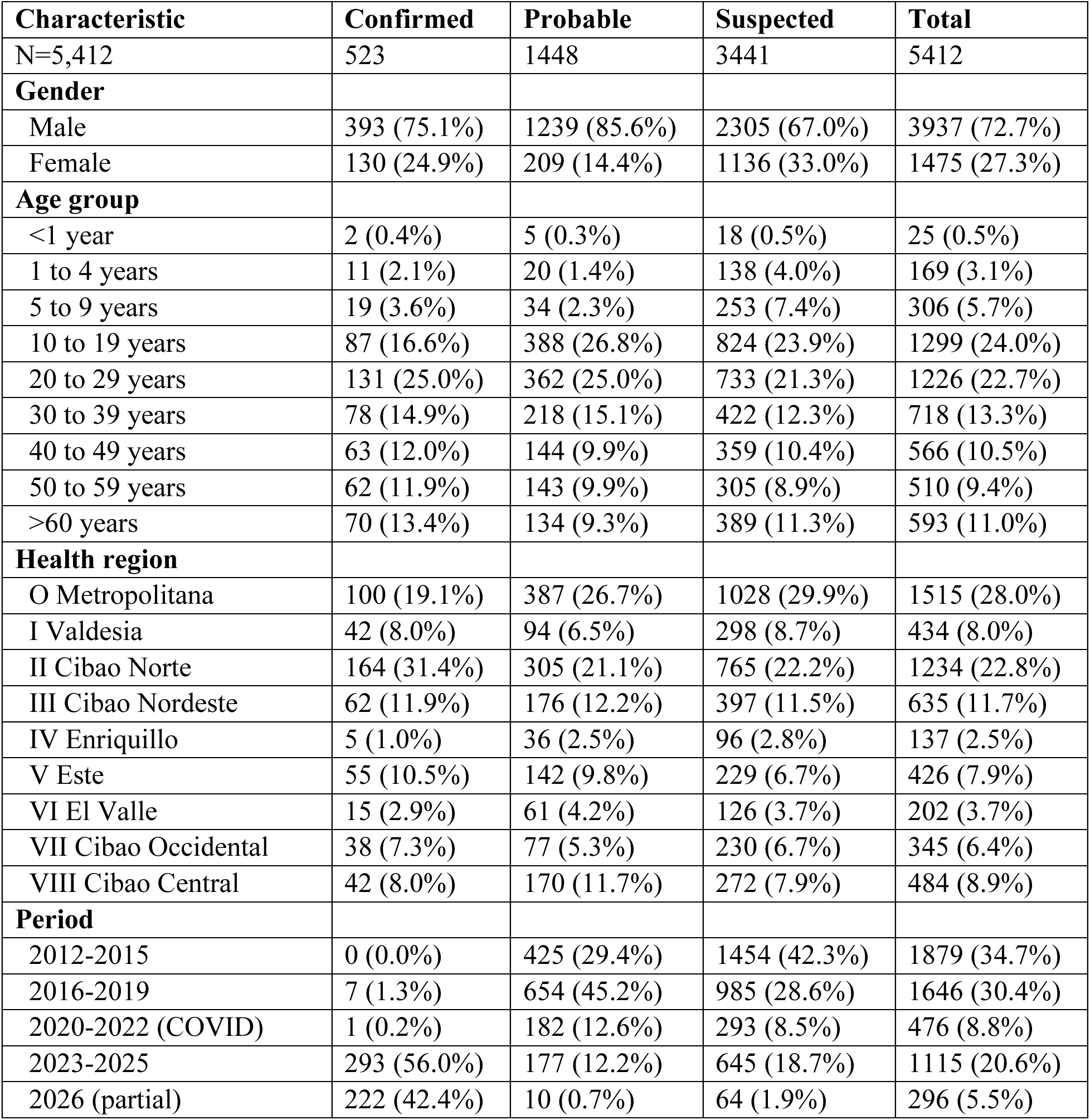
Sociodemographic and clinical characteristics of leptospirosis cases (N=5,412), stratified by case classification, 2012–2026.

| **Characteristic** | **Confirmed** | **Probable** | **Suspected** | **Total** |
| --- | --- | --- | --- | --- |
| N=5,412 | 523 | 1448 | 3441 | 5412 |
| **Gender** |  |  |  |  |
| Male | 393 (75.1%) | 1239 (85.6%) | 2305 (67.0%) | 3937 (72.7%) |
| Female | 130 (24.9%) | 209 (14.4%) | 1136 (33.0%) | 1475 (27.3%) |
| **Age group** |  |  |  |  |
| <1 year | 2 (0.4%) | 5 (0.3%) | 18 (0.5%) | 25 (0.5%) |
| 1 to 4 years | 11 (2.1%) | 20 (1.4%) | 138 (4.0%) | 169 (3.1%) |
| 5 to 9 years | 19 (3.6%) | 34 (2.3%) | 253 (7.4%) | 306 (5.7%) |
| 10 to 19 years | 87 (16.6%) | 388 (26.8%) | 824 (23.9%) | 1299 (24.0%) |
| 20 to 29 years | 131 (25.0%) | 362 (25.0%) | 733 (21.3%) | 1226 (22.7%) |
| 30 to 39 years | 78 (14.9%) | 218 (15.1%) | 422 (12.3%) | 718 (13.3%) |
| 40 to 49 years | 63 (12.0%) | 144 (9.9%) | 359 (10.4%) | 566 (10.5%) |
| 50 to 59 years | 62 (11.9%) | 143 (9.9%) | 305 (8.9%) | 510 (9.4%) |
| >60 years | 70 (13.4%) | 134 (9.3%) | 389 (11.3%) | 593 (11.0%) |
| **Health region** |  |  |  |  |
| O Metropolitana | 100 (19.1%) | 387 (26.7%) | 1028 (29.9%) | 1515 (28.0%) |
| I Valdesia | 42 (8.0%) | 94 (6.5%) | 298 (8.7%) | 434 (8.0%) |
| II Cibao Norte | 164 (31.4%) | 305 (21.1%) | 765 (22.2%) | 1234 (22.8%) |
| III Cibao Nordeste | 62 (11.9%) | 176 (12.2%) | 397 (11.5%) | 635 (11.7%) |
| IV Enriquillo | 5 (1.0%) | 36 (2.5%) | 96 (2.8%) | 137 (2.5%) |
| V Este | 55 (10.5%) | 142 (9.8%) | 229 (6.7%) | 426 (7.9%) |
| VI El Valle | 15 (2.9%) | 61 (4.2%) | 126 (3.7%) | 202 (3.7%) |
| VII Cibao Occidental | 38 (7.3%) | 77 (5.3%) | 230 (6.7%) | 345 (6.4%) |
| VIII Cibao Central | 42 (8.0%) | 170 (11.7%) | 272 (7.9%) | 484 (8.9%) |
| **Period** |  |  |  |  |
| 2012-2015 | 0 (0.0%) | 425 (29.4%) | 1454 (42.3%) | 1879 (34.7%) |
| 2016-2019 | 7 (1.3%) | 654 (45.2%) | 985 (28.6%) | 1646 (30.4%) |
| 2020-2022 (COVID) | 1 (0.2%) | 182 (12.6%) | 293 (8.5%) | 476 (8.8%) |
| 2023-2025 | 293 (56.0%) | 177 (12.2%) | 645 (18.7%) | 1115 (20.6%) |
| 2026 (partial) | 222 (42.4%) | 10 (0.7%) | 64 (1.9%) | 296 (5.5%) |

Among the 5,412 cases included in our study, 3,441 (63.6 %) were classified as suspected, 1,448 (26.8 %) as probable, and 523 (9.7 %) as confirmed. The cohort was predominantly male (72.7 %), a proportion that was even higher among probable (85.6 %) and confirmed (75.1 %) cases than among suspected cases (67.0 %). The Metropolitana region (Zone 0) accounted for the largest share of cases (28.0 %), followed by II Cibao Norte (22.8 %). No confirmed case was recorded between 2012 and 2015, and only 1.3 % of confirmed cases occurred between 2016 and 2019. In contrast, 56.0 % of confirmed cases were concentrated in 2023–2025, and a further 42.4 % occurred in 2026 alone, over a six-month observation window (Figure 1).

**Figure 1.**
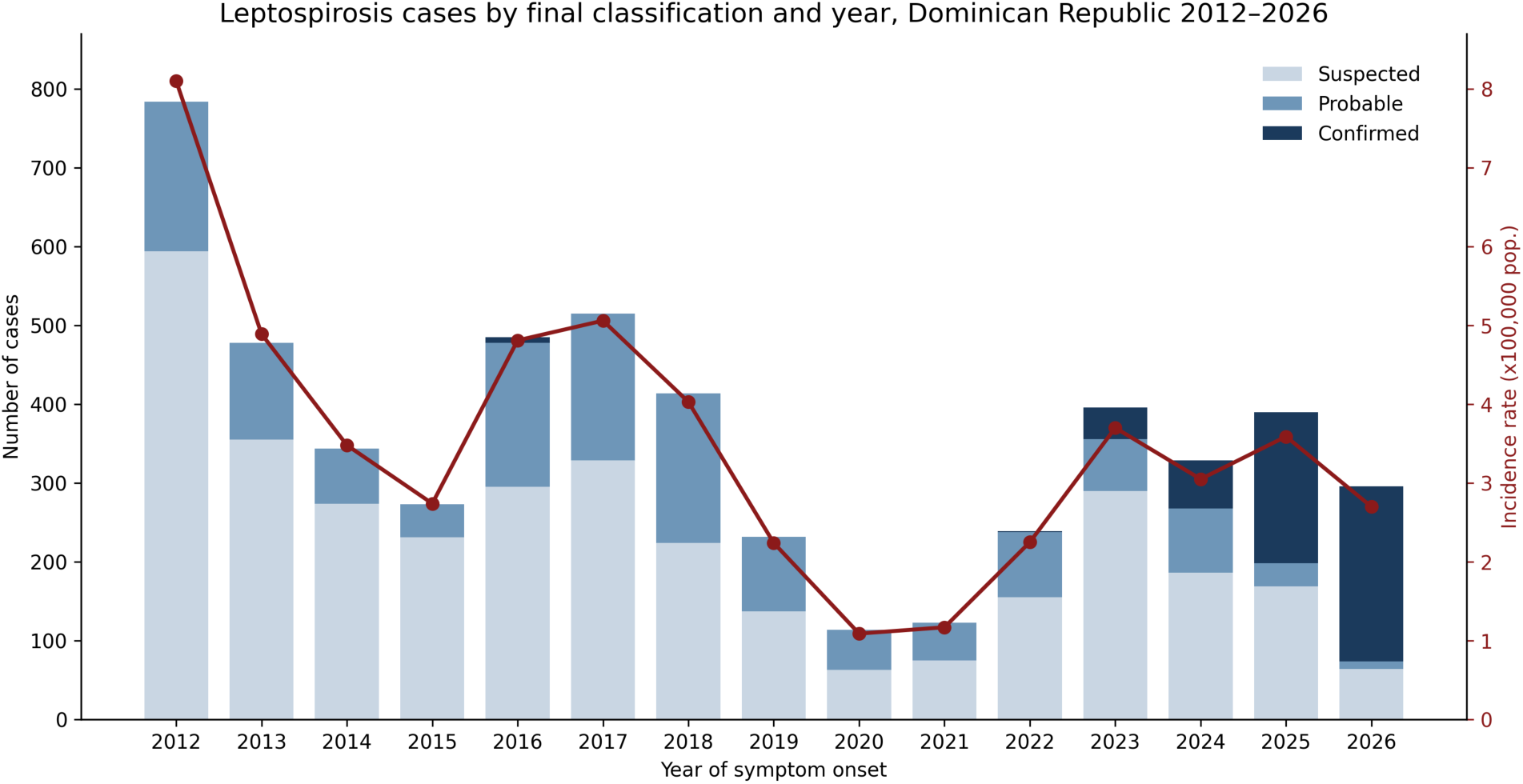
Annual epidemic curve by final case classification, with overall incidence rate overlaid, Dominican Republic, 2012-2026 (June)

### Age and gender distribution

Incidence was considerably higher in men than in women throughout the study period (male:female ratio = 2.73; 95% CI 2.56–2.90). This male predominance was evident across all case classifications but became more pronounced with increasing diagnostic certainty: men accounted for 67.0% of suspected cases, rising to 82.8% of cases classified as probable or confirmed (85.6% and 75.1%, respectively; Table 1). By age group, incidence in men peaked between 10 and 29 years (approximately 7 cases per 100,000 person-years), a pattern reflected in the raw case counts, where the 10–19 and 20–29-year age bands together accounted for 46.7% of all cases (24.0% and 22.7%, respectively; Table 1). In contrast, incidence in women remained low across all age groups, ranging from 0.54 (age <1 year) to 2.47 (age 20–29 years) cases per 100,000 person-years, without an equivalent peak in young adulthood (Table 2). Beyond the youngest age groups, both confirmed and probable cases showed a gradual decline with advancing age through the 40– 49 and 50–59-year bands, followed by a modest uptick in the oldest group (>60 years, 13.4% of confirmed and 11.0% of all cases; Table 1), a pattern that did not mirror the more consistent decline seen among suspected cases. Incidence rates in Table 2 (N=5,116) exclude the 296 partial-year 2026 cases included in the case-classification totals in Table 1 (N=5,412), reflecting the exclusion of an incomplete person-years denominator for 2026.

**Table 2.** Age- and sex-specific leptospirosis incidence rates per 100,000 person-years, 2012– 2025 (excludes partial-year 2026 data)

| Age group | Gender | Cases | Person-years | Rate x100,000 | 95% CI |
| --- | --- | --- | --- | --- | --- |
| <1 year | Male | 16 | 1,367,490 | 1.17 | 0.67–1.90 |
|  | Female | 7 | 1,304,766 | 0.54 | 0.22–1.11 |
|  | **Total** | **23** | **2,672,256** | **0.86** | **0.55–1.29** |
| 1 to 4 years | Male | 82 | 5,466,266 | 1.50 | 1.19–1.86 |
|  | Female | 67 | 5,250,404 | 1.28 | 0.99–1.62 |
|  | **Total** | **149** | **10,716,670** | **1.39** | **1.18–1.63** |
| 5 to 9 years | Male | 170 | 6,886,266 | 2.47 | 2.11–2.87 |
|  | Female | 121 | 6,627,753 | 1.83 | 1.51–2.18 |
|  | **Total** | **291** | **13,514,019** | **2.15** | **1.91–2.42** |
| 10 to 19 years | Male | 935 | 13,539,578 | 6.91 | 6.47–7.36 |
|  | Female | 313 | 13,208,523 | 2.37 | 2.11–2.65 |
|  | **Total** | **1,248** | **26,748,101** | **4.67** | **4.41–4.93** |
| 20 to 29 years | Male | 861 | 12,280,392 | 7.01 | 6.55–7.50 |
|  | Female | 304 | 12,285,287 | 2.47 | 2.20–2.77 |
|  | **Total** | **1,165** | **24,565,679** | **4.74** | **4.47–5.02** |
| 30 to 39 years | Male | 503 | 10,255,983 | 4.90 | 4.49–5.35 |
|  | Female | 177 | 10,447,197 | 1.69 | 1.45–1.96 |
|  | **Total** | **680** | **20,703,180** | **3.28** | **3.04–3.54** |
| 40 to 49 years | Male | 411 | 8,342,791 | 4.93 | 4.46–5.43 |
|  | Female | 123 | 8,527,268 | 1.44 | 1.20–1.72 |
|  | **Total** | **534** | **16,870,059** | **3.17** | **2.90–3.45** |
| 50 to 59 years | Male | 348 | 6,486,604 | 5.36 | 4.82–5.96 |
|  | Female | 128 | 6,647,582 | 1.93 | 1.61–2.29 |
|  | **Total** | **476** | **13,134,186** | **3.62** | **3.31–3.96** |
| >60 years | Male | 415 | 7,407,367 | 5.6 | 5.08–6.17 |
|  | Female | 135 | 7,857,612 | 1.72 | 1.44–2.03 |
|  | **Total** | **550** | **15,264,979** | **3.6** | **3.31–3.92** |
| **All ages** | Male | 3,741 | 72,032,737 | 5.19 | 5.03–5.36 |
|  | Female | 1,375 | 72,156,392 | 1.91 | 1.81–2.01 |
|  | **Total** | **5,116** | **144,189,129** | **3.55** | **3.45–3.65** |

### Temporal Trends

The total national incidence rate declined from 8.10 per 100,000 population in 2012 to a low 2.74 in 2015, and remained relatively stable, between 2 and 4 per 100,000, for most of the following decade (Figure 1). The trend analysis showed no significant annual percent change in the total number of cases over the 2012–2025 period (p = 0.15).

The rate of confirmed cases, by contrast, followed a markedly different trajectory: it remained at zero until 2015, stayed marginal through 2022, and rose steadily from 2023 onward, reaching 1.77 per 100,000 population in 2025 and 2.03 in the first half of 2026 (not yet annualized). This increase corresponded to an annual percent change of +42.6 % (p = 0.007), markedly different from the flat trend observed for total cases.

The interrupted time-series model of monthly cases, taking COVID-19 onset as an interruption event (Figure 2), showed a gradual downward trend from 2012 to early 2020, declining from approximately 52 to 24 cases per month despite an extreme outlier spike near 150 cases in 2017. At COVID onset, the fitted trend dropped sharply to approximately 5 cases per month before climbing steadily back to approximately 25 cases per month by 2023. A second level shift occurred at the 2023 resurgence onset, with the trend rising to approximately 31 cases per month and plateauing through 2025.

**Figure 2.**
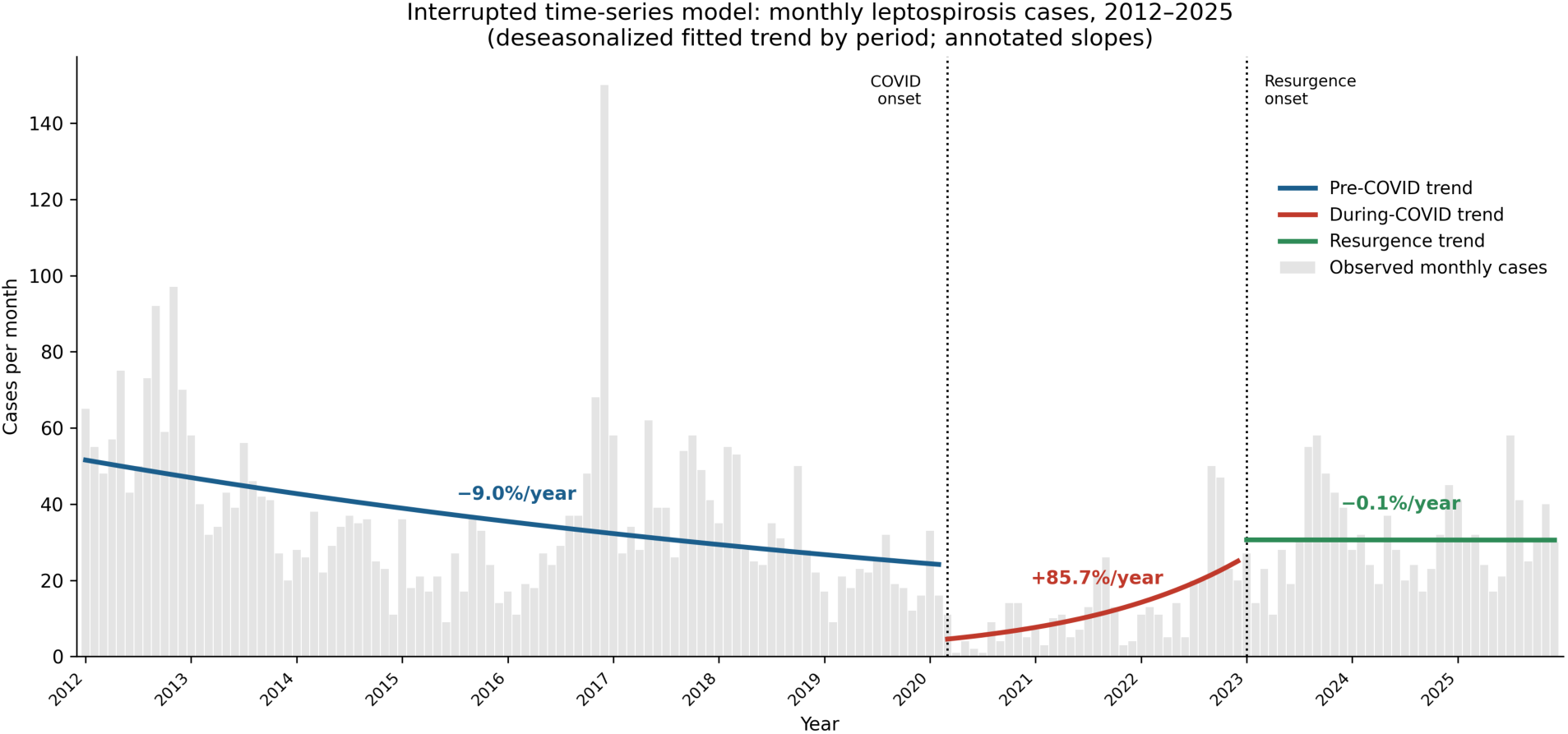
Interrupted time-series analysis of monthly case counts, 2012–2025, Dominican Republic, 2012-2025.

### Seasonal Trends

We found a marked seasonal pattern in the monthly distribution of cases (chi-square goodness-of-fit test, p < 0.001), with 61.1 % of cases concentrated in the rainy/hurricane season (May– November) versus 38.9 % in the dry season (Figure 3). Monthly rainfall climatology showed a moderate positive correlation with this seasonality (r = 0.553), though of marginal significance given the small number of monthly observations (p = 0.062) (Figure 4).

**Figure 3.**
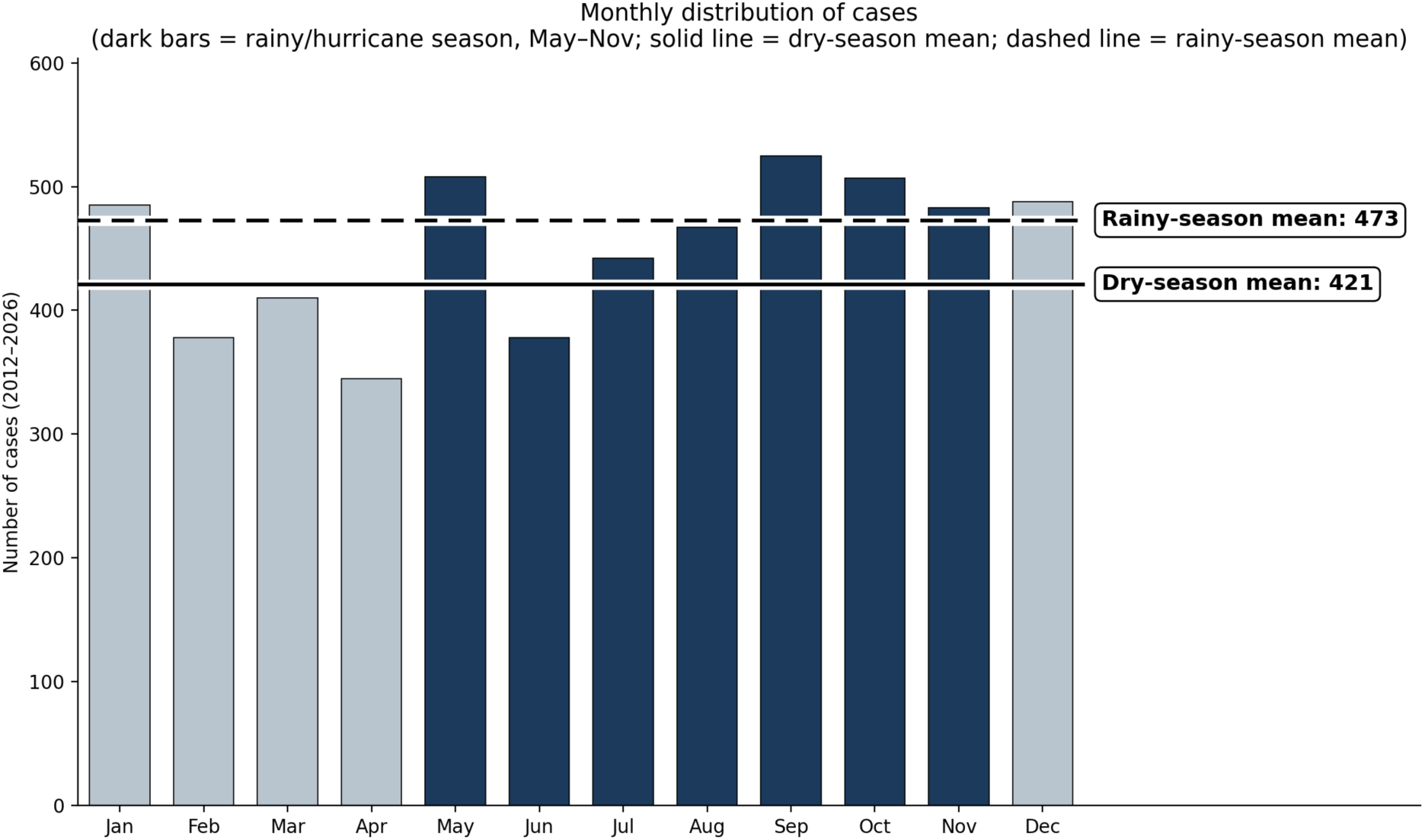
Seasonal distribution of cases by month, Dominican Republic, 2012-2026.

**Figure 4.**
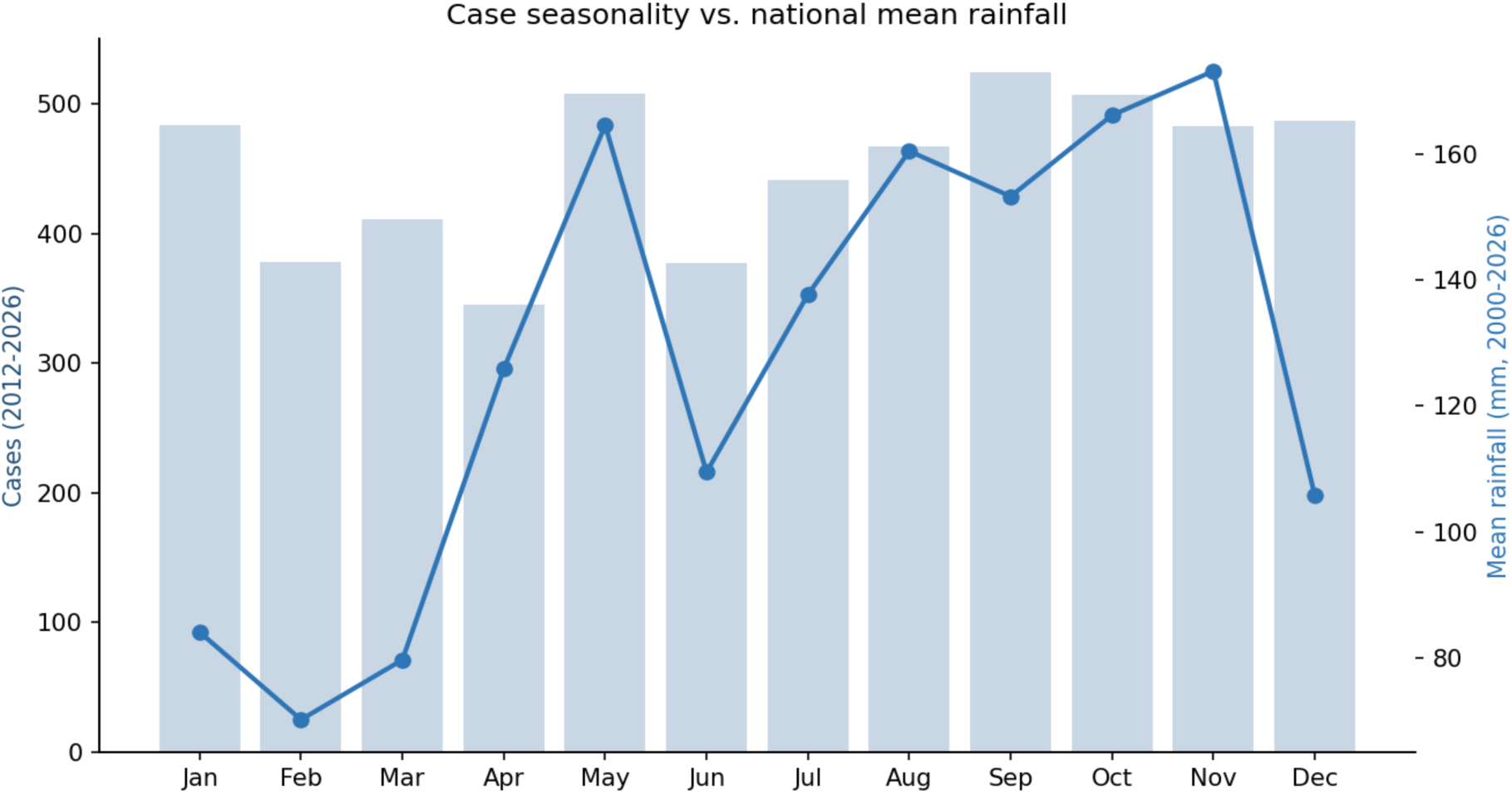
Seasonal distribution of cases relative to national mean rainfall, Dominican Republic, 2012-2026.

### Geographic distribution

The provinces with the highest average incidence rate over the 2012–2025 period were Hermanas Mirabal (16.43 per 100,000 person-years), San José de Ocoa (11.26), Monte Cristi (10.53), Espaillat (9.66), and Duarte (5.90) (Table 3). This ranking contrasted sharply with that based on the absolute number of cases, led by Santo Domingo, Santiago, and the National District, provinces that, despite concentrating the largest case volume, ranked mid- to low once adjusted for population size. At the health-region level, III Cibao Nordeste (6.79 per 100,000 person-years) and VII Cibao Occidental (5.69) recorded the highest rates in the country, while the O Metropolitana region —which accounts for the largest share of cases in absolute terms, as noted above— showed one of the lowest rates (2.58), only above IV Enriquillo (2.49) (data not shown).

**Table 3.**
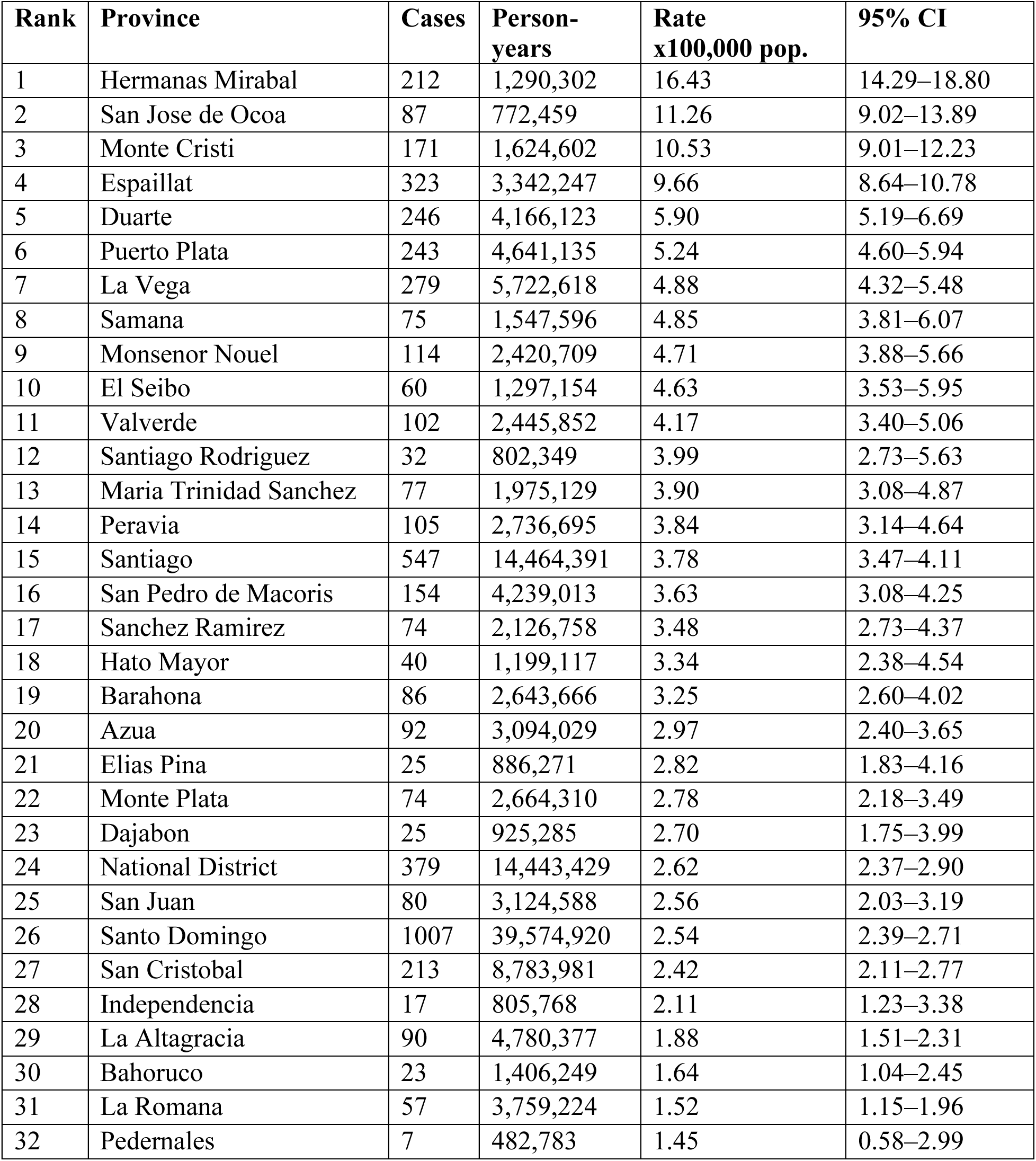
Leptospirosis incidence rates by province, ranked from highest to lowest, 2012– 2025.

When incidence rates aggregated into eight geographic zones (Table 4), the semi-arid Northwest (Monte Cristi, Dajabon) has the highest rate at 7.69 per 100,000 (95% CI: 6.65–8.84), followed by the Cibao Valley (5.24; 5.01–5.48) and North Atlantic coast (4.84; 4.37–5.34), while the Southwest arid/Enriquillo basin has the lowest rate at 2.49 (2.09–2.95) despite also being classified as arid. These data do not support the hypothesis of a simple “aridity drives risk” narrative. This divergence between the two arid zones likely reflects local hydrology and land use. Specifically, the Northwest’s elevated rate appears driven by Monte Cristi’s irrigated agriculture along the Yaque del Norte river, which sustains animal reservoirs and standing-water exposure, whereas the more desertic, less-irrigated Enriquillo basin lacks these conditions despite similar aridity. The Metropolitan zone and Southeast plains sit near the bottom (2.57, 2.43–2.70; and 2.65, 2.42–2.90) despite dense population and high case counts, showing that urbanization does not straightforwardly predict risk once normalized. Since this classification is qualitative and narratively assigned rather than GIS-derived, confirming these hydrology- and land-use-based drivers would require formal analysis linking province-level covariates (irrigation extent, livestock density, land cover) to the surveillance data.

**Table 4.** Leptospirosis incidence rates by geographic zone, 2012–2025.

| **Geographic zone** | **Provinces**<br>**(n)** | **Cases** | **Rate**<br>**x100,000** | **95% CI** |
| --- | --- | --- | --- | --- |
| Semi-arid Northwest | 2 | 196 | 7.69 | 6.65–8.84 |
| Cibao Valley (agricultural interior) | 9 | 1929 | 5.24 | 5.01–5.48 |
| North Atlantic coast | 3 | 395 | 4.84 | 4.37–5.34 |
| Central mountains (Cordillera Central) | 3 | 192 | 4.01 | 3.47–4.62 |
| South-central plains/coast | 3 | 410 | 2.81 | 2.54–3.09 |
| Southeast plains | 6 | 475 | 2.65 | 2.42–2.90 |
| Metropolitan (Santo Domingo) | 2 | 1386 | 2.57 | 2.43–2.70 |
| Southwest arid / Enriquillo basin | 4 | 133 | 2.49 | 2.09–2.95 |

### Clinical predictors of complications

In the multivariable logistic regression model (Table 5), pre-existing comorbidity was the strongest predictor of clinical complications (odds ratio [OR] = 2.84; 95 % CI 2.01–4.02; p < 0.001). Calendar year was positively and significantly associated with the probability of complications (OR = 1.07 per year; p = 0.004), consistent with the increase in clinical documentation observed in recent years. Gender was not independently associated with complications (OR = 0.99; p = 0.97) after adjusting for age and comorbidity. The 5–9-year (OR = 0.32; p = 0.015), 10–19-year (OR = 0.46; p = 0.002), and over-60 (OR = 0.58; p = 0.031) age groups showed significantly lower odds of complications than the reference group (20–29 years).

**Table 5.** Multivariable logistic regression of predictors of clinical complications.

| **Variable** | **OR** | **95% CI**<br>**low** | **95% CI**<br>**high** | **p-value** |
| --- | --- | --- | --- | --- |
| Intercept | 0.31 | 0.20 | 0.48 | <0.0001 |
| Male gender (ref: female) | 0.99 | 0.70 | 1.42 | 0.971 |
| Comorbidity present (ref: absent) | 2.84 | 2.01 | 4.02 | <0.0001 |
| Year (centered) | 1.07 | 1.02 | 1.12 | 0.0036 |
| Age <1 year (ref: 20-29) | 0.59 | 0.06 | 5.43 | 0.6436 |
| Age 1-4 years (ref: 20-29) | 0.86 | 0.25 | 2.89 | 0.8026 |
| Age 5-9 years (ref: 20-29) | 0.32 | 0.13 | 0.80 | 0.0154 |
| Age 10-19 years (ref: 20-29) | 0.46 | 0.28 | 0.75 | 0.0019 |
| Age 30-39 years (ref: 20-29) | 0.83 | 0.51 | 1.35 | 0.4485 |
| Age 40-49 years (ref: 20-29) | 0.76 | 0.45 | 1.28 | 0.3011 |
| Age 50-59 years (ref: 20-29) | 0.80 | 0.47 | 1.36 | 0.4201 |
| Age >60 years (ref: 20-29) | 0.58 | 0.35 | 0.95 | 0.0309 |

Taken together, these findings indicate that the overall burden of disease remained relatively stable from 2012 to 2025, while the diagnostic profile shifted substantially in the most recent period, marked by a significant increase in confirmed cases beginning in 2023. Incidence varied geographically and did not necessarily correspond to areas with the highest absolute case counts. Cases were consistently concentrated among young and middle-aged men, with a clear seasonal pattern associated with the rainy season. Among clinical factors, pre-existing comorbidities showed the strongest association with a complicated disease course, highlighting an important subgroup for clinical risk assessment and management.

## Discussion

Differential association of sociodemographic, geographic, and environmental factors with leptospirosis in the Dominican Republic suggests a tailored control strategy that aims to reduce the incidence of disease while strengthening the capacity for diagnostic testing and screening. The noticeable gender differences, along with the concentration of male risk in young adulthood, confirms occupational and behavioral exposure patterns documented internationally. These include agricultural work, outdoor labor, and contact with contaminated water or soil, all of which, disproportionately affect young and middle-aged men across most settings where leptospirosis has been studied.^10,11,14,16,25,46^ This pattern is also consistent with reports that young adults face elevated risk through recreational, school-related, and travel-associated water exposure, in addition to occupational contact, with risk declining at older ages as exposure diminishes.^14,15^ The fact that the male skew persisted across suspected, probable, and confirmed cases alike argues against this simply reflecting broader case ascertainment, and points instead to exposure tied to the life stage at which young men typically enter manual labor or spend more time in recreational water. The uniformly low incidence in women likely reflects lower-risk domestic or peridomestic exposures compared to the more direct occupational and environmental contact typical of young men.^10,11,16^

This interpretation should still be considered alongside surveillance limitations, since differential healthcare access by gender could inflate the observed ratio somewhat. Still, the persistence of the male skew from suspected to more diagnostically certain cases, together with its alignment with international estimates,^10,11,14,16,25,46^ makes a purely artifactual explanation unlikely, though the skew is somewhat attenuated among the smaller subset of cases reaching laboratory confirmation. Moreover, the modest rise in cases among adults over 60 also stands out and warrants further investigation, as it could reflect a distinct exposure or susceptibility pathway not captured by the occupational hypothesis. Disentangling this would require data, such as occupation, comorbidities, rural or urban residence, that are not currently linked to the surveillance dataset.

Another remarkable finding is the divergence between a stable overall incidence rate across 2012–2025 and a sharply rising rate of confirmed cases from 2023 onward. To evaluate whether pandemic era shifts in reported incidence reflected true changes in transmission or artifacts of disrupted healthcare seeking behavior and diagnostic capacity, we examined the interrupted time-series pattern in more detail. The gradual pre-pandemic decline is consistent with a modest, real decline in transmission or improved control over the decade preceding COVID. However, the abrupt drop to ∼5 cases per month at COVID onset is too sudden to reflect genuine epidemiological change, and is more consistent with disrupted healthcare-seeking behavior, reduced surveillance capacity, and diagnostic resources being redirected to the pandemic response.^47,48^ This pattern has been previously documented for leptospirosis in Sri Lanka, where the country’s largest-ever outbreak in 2020 went largely uninvestigated as health system attention shifted to COVID-19,^49^ and in Thailand, where surveillance disruption and underreporting were similarly implicated in reduced case detection during the pandemic.^15^

The steady recovery back to ∼25 cases per month by 2023 as these disruptions eased supports this interpretation, mirroring the dip-and-resurgence pattern observed for other notifiable diseases such as sexually transmitted infections in the United States following pandemic-related staffing and resource diversion.^50^ The subsequent plateau through 2025, below late pre-COVID peaks but above the COVID-era trough, suggests that reported incidence stabilized at a new, moderately elevated baseline. Together, these findings suggest the pandemic-era pattern reflects a temporary collapse and subsequent recovery of surveillance and reporting infrastructure rather than a genuine new disease surge,^47,48^ with true underlying transmission likely closer to the gently declining pre-COVID trend than either the artificially low COVID-era estimate or the post-resurgence plateau would suggest in isolation.

We interpret these patterns cautiously. The proportion of cases reaching laboratory confirmation increased substantially from the early study period to 2025–2026, a shift that plausibly reflects expanded diagnostic capacity, broader access to PCR or culture-based confirmation, or strengthened laboratory referral pathways, rather than a genuine increase in *Leptospira* circulation. This distinction matters for how the finding should be used: a rising confirmed-case rate driven by diagnostics signals a maturing surveillance system, while the same rate driven by transmission would signal an emerging public health warning requiring a different response.

This diagnostic uncertainty is not unique to our setting. It is a longstanding feature of leptospirosis surveillance globally, reflecting the technical demands and limited availability of confirmatory serological and molecular assays,^8^ and our data are consistent with that broader pattern. Confirmed-case counts should therefore be interpreted as a conservative estimate of true disease burden, consistent with the Dominican Republic’s national seroepidemiologic survey showing that reported cases capture only a small fraction of actual infections.^13^ Both suspected and confirmed cases likely underestimate underlying pathogen circulation.

This study extends a small but growing body of evidence from research on leptospirosis in the Dominican Republic. In two recent analyses examined environmental and sociodemographic drivers of transmission at the subnational level: a geospatial cluster analysis identified high-risk areas in two provinces,^43^ and a geographically weighted regression modeling how these drivers vary in strength across space.^44^ Our national, 14-year dataset allows us to address whether the clustering patterns identified in those more geographically limited studies hold at scale. The provinces with the highest population-adjusted incidence in our cohort (Hermanas Mirabal, San José de Ocoa, Monte Cristi, Espaillat, and Duarte) offer a natural point of comparison to the clusters previously reported,^43,44^ a formal overlap analysis was beyond the scope of this study, but if future work confirms alignment between these provinces and the previously reported clusters, it would strengthen the case that they are durable, structural sources of risk rather than artifacts of a particular study period or sampling frame.

The geographic pattern suggests a more specific mechanism than aridity alone. Both the highest- and lowest-incidence zones were classified as arid, suggesting that differences in water availability and use within these climate zones may help explain the observed variation. Irrigated agriculture along the Yaque del Norte may sustain standing water and animal reservoirs that support leptospirosis transmission, even in otherwise dry settings. However, because the zone classifications were based on narrative descriptions rather than validated GIS-derived land-cover or elevation data, these findings should be interpreted cautiously. Future analyses linking local irrigation, land use, terrain, and surveillance data will be needed to determine whether these factors explain the geographic differences in incidence.

Another interesting trend is the disconnect between where cases occur in absolute numbers and where population-adjusted risk is highest. Santo Domingo, Santiago, and the National District accounted for the largest case volumes, consistent with their large populations, but ranked mid- to low once incidence was adjusted for population size. Conversely, smaller and more rural provinces carried the highest per-capita burden. This pattern was also apparent in findings from Brazil, where urban environments and rural agricultural settings each generate distinct transmission dynamics that are obscured when data were examined only in absolute terms.^12,51^ For a national surveillance system, this has a direct programmatic implication: resource allocation and targeted interventions based on raw case counts alone would systematically under-prioritize the smaller provinces where an individual’s risk of infection is in fact highest.

The seasonal concentration of cases during the rainy/hurricane season (May–November) is consistent with the well-established role of rainfall and flooding in facilitating environmental *Leptospira* transmission.^10,16,21^ However, the monthly correlation with rainfall climatology in our data was only of marginal statistical significance, likely reflecting the limited statistical power of month-level aggregation over a single climatological cycle. Studies using finer temporal resolution and explicitly modeling lagged rainfall effects have found stronger and more clearly delineated relationships between precipitation, flooding, and case onset, with lags often spanning several weeks between rainfall events and clinical presentation.^17,18,19^ Similarly, hydroclimatic modeling approaches applied elsewhere in Latin America have captured non-linear and threshold effects that a simple monthly correlation cannot detect.^20,33,34^ Future work using disaggregated, lagged rainfall data — potentially integrated into an early warning framework analogous to that developed for northeastern Argentina^52^ — could substantially sharpen the environmental signal identified here and translate it into an operational forecasting tool for the Ministry of Public Health.

In our multivariable model, pre-existing comorbidity was the strongest independent predictor of clinical complications, while the positive association between calendar year and complication risk plausibly reflects the same improvement in clinical documentation and diagnostic completeness that underlies the rising confirmed-case trend discussed above, rather than a true increase in disease severity over time. This pattern is broadly consistent with clinical reviews in which comorbid conditions were key determinants of severe leptospirosis outcomes^5,6^ and it reinforces the interpretation that changes in what the surveillance system captures — not necessarily changes in the underlying disease — explain much of the temporal signal in this dataset.

Several limitations should be considered. First, a substantial proportion of records were excluded due to pending or missing classification, which may bias the cohort if missingness is non-random with respect to severity, geography, or time. Second, as a passive system, underreporting is likely substantial and uneven, consistent with the low reporting fraction identified in the national seroepidemiologic survey.^13^ Third, the rising proportion of confirmed cases over time raises the possibility of differential misclassification bias, complicating comparisons of severity or demographics across years. Fourth, our reliance on provincial-level meteorological aggregates precludes the fine-scale resolution achieved by direct environmental sampling, which has shown that transmission risk can vary sharply even within a single neighborhood.^23,53–55^ Finally, our data do not distinguish exposure route: international tourism is a distinct, unmeasured pathway, as travel-acquired leptospirosis has been linked to freshwater recreation in Southeast Asia, the Caribbean, and Latin America.^42^ Since the United States alone accounts for roughly half of the Dominican Republic’s international arrivals,^56^ travel-associated exposure among visitors may be an underrecognized transmission route alongside the occupational and environmental exposures captured by routine surveillance.^42,56^

Despite these limitations, this study provides the Ministry of Public Health with the first systematic, multi-decade situational analysis of leptospirosis in the Dominican Republic. We have identified priority provinces, at-risk demographic groups, and a seasonal window for targeted intervention. A central finding with direct programmatic relevance is the persistent gap between suspected and confirmed diagnosis: laboratory confirmation was virtually absent before 2016 and reached over 40% of cases only from 2023 onward, indicating that for most of the study period the surveillance system was structurally limited in its capacity to confirm leptospirosis early in the clinical course. This gap had direct consequences for timely treatment and case management.^5,6^

The integration of geographically disaggregated meteorological data further strengthened this analysis by enabling a more granular understanding of how rainfall and flooding patterns relate to disease risk at the provincial level,^21,44^ findings that are directly actionable for early warning systems,^52^ targeted environmental interventions, and climate-adaptive public health planning.^57^ Taken together, these findings align with and extend the objectives of the Global Leptospirosis Environmental Action Network,^58^ respond to calls for stronger national-level surveillance data raised in prior Caribbean-wide reviews.^40,41^ They also directly inform the Ministry’s capacity to assess its surveillance infrastructure, evaluate existing vector and environmental control measures, and develop targeted, evidence-based interventions. Strengthening early diagnostic capacity, alongside translating the geographic and seasonal patterns identified here into an operational early warning and resource allocation framework, represents a logical next step for the country’s response to this persistently neglected disease.

## Conclusion

Leptospirosis remains a neglected tropical disease with substantial morbidity in resource-limited settings, despite lacking formal recognition on the WHO’s Neglected Tropical Disease list.^1,7^ This study fills a longstanding evidence gap by providing the first systematic, multi-decade situational analysis of leptospirosis in the Dominican Republic, building on the country’s only national seroepidemiologic survey, which noted how little was known about local disease epidemiology and how substantially reported cases underestimate true infection burden.^13^

Confirmatory diagnosis emerged as a particular challenge throughout the surveillance period, with laboratory-confirmed cases virtually absent before 2016 and exceeding 40% of the total only from 2023 onward, highlighting the need for sustained investment in diagnostic infrastructure to support timely case detection and management.^5,6,8^ By identifying high-risk provinces, at-risk demographic groups, seasonal transmission windows, and, critically, a persistent gap in early diagnostic confirmation, these findings offer an evidence base for strengthening national surveillance, diagnostic capacity, and climate-adaptive prevention strategies, while contributing to regional efforts such as the Global Leptospirosis Environmental Action Network^58^ to address leptospirosis in tropical island and Caribbean settings.^40,41^ Given the country’s major tourism sector, these findings may also inform traveler health guidance and clinician awareness, representing a distinct exposure pathway from the primarily occupational and environmental drivers described here.^42,56^

## Authors’ Contributions

Conceptualization, L.V.A., J.J.S., and T.D.D.; methodology, L.V.A. and J.J.S.; validation, L.V.A., J.J.S., and O.A.A.; formal analysis, J.J.S. and L.V.A.; investigation, L.V.A., D.D.L.; H.A.S., and L.S.; resources, J.L.C.; data curation, J.J.S.; writing—original draft preparation, L.V.A.; writing—review and editing. J.J.S., O.A.A., B.L.M., C.R.M., C.H.H., and T.D.D.; supervision, T.D.D. All authors have read and agreed to the published version of the manuscript.

## Ethics Declarations

This study was reviewed by the Florida Atlantic University Institutional Review Board (protocol IRB2606182, “Leptospirosis in the Dominican Republic, 2000–2026: National Surveillance Trends”), which determined it exempt from federal regulation under 45 CFR 46.104(4)(ii) on June 22, 2026, as a secondary analysis of de-identified national surveillance and meteorological data. Individual informed consent was not required, consistent with this exemption determination.

## Data Availability Statement

The surveillance dataset analyzed in this study was provided by the Dominican Republic Ministry of Public Health and Social Assistance (MISPAS) through the General Directorate of Epidemiology (DIGEPI). Precipitation data were obtained from the Dominican Institute of Meteorology (INDOMET). Because these data are derived from national surveillance and meteorological systems and may contain sensitive information, they are not publicly available. De-identified data may be requested from the corresponding author or directly from DIGEPI/MISPAS and INDOMET, subject to a reasonable request and approval by the relevant institutions.

## Financial Support

This research was not funded by any agency in the public, commercial, or not-for-profit sectors.

## Acknowledgments

We thank the General Directorate of Epidemiology (DIGEPI) of the Dominican Republic’s Ministry of Public Health and the Dominican Institute of Meteorology (INDOMET) for providing access to the national surveillance and rainfall data that made this study possible.

## Conflict of Interests

The authors declare that they have no competing interests. Professor Hennekens also declares that he serves as an independent scientist in an advisory role to investigators and sponsors as Chair of two Data Moni-toring Committees for Amgen (erenumab and evolocumab); to the United States Food and Drug Administration as a Special Government Employee (SGE) and UpToDate; receives royalties for authorship or editorship of 3 textbooks; has an investment management relationship with the West-Bacon Group within Truist Investment Services, which has discretionary investment authority; does not own any common or preferred stock in any pharmaceutical or medical device company.

## Declaration of Generative AI and AI-Assisted Technologies

During the preparation of this manuscript, the authors used Claude as a language-editing tool to improve clarity, readability, and overall presentation. AI assistance was limited to language and editorial support and was not used for data analysis, statistical modeling, interpretation of findings, or development of scientific conclusions. The authors independently reviewed and approved all AI-assisted text and remain fully responsible for the accuracy, integrity, and final content of the manuscript.

## Current Contact Addresses

Lisette V. Alcántara, Florida Atlantic University, Boca Raton, FL, United States.

Jose J. Sánchez, Pontificia Universidad Católica Madre y Maestra, Santiago, Dominican Republic.

Oscar A. Aleuy, Florida Atlantic University, Boca Raton, FL, United States.

David De Luna, Pontificia Universidad Católica Madre y Maestra, Santiago, Dominican Republic.

Benjamin L. Miller, University of Rochester, Rochester, NY, United States.

Charles R. Mace, Tufts University, Medford, MA, United States.

Hunter Scott, Florida Atlantic University, Boca Raton, FL, United States.

Lara Smejkal, Florida Atlantic University, Boca Raton, FL, United States.

Jose L. Cruz, Ministry of Public Health, Santo Domingo, Dominican Republic.

Charles H. Hennekens, Florida Atlantic University, Boca Raton, FL, United States.

Timothy D. Dye, Florida Atlantic University, Boca Raton, FL, United States.

## List Of Abbreviations

### Abbreviation Definition

APC: Annual percent change
CECOVEZ: Center for the Prevention and Control of Vector-Borne and Zoonotic Diseases
CI: Confidence interval
COVID-19: Coronavirus disease 2019
DALYs: Disability-adjusted life years
DIGEPI: General Directorate of Epidemiology (Dirección General de Epidemiología)
INDOMET: Dominican Institute of Meteorology (Instituto Dominicano de Meteorología)
IRB: Institutional Review Board
IRLS: Iteratively reweighted least squares
MAT: Microscopic agglutination testing
MISPAS: Ministry of Public Health and Social Assistance
OR: Odds ratio
PCR: Polymerase chain reaction
PUCMM: Pontificia Universidad Católica Madre y Maestra
SNS: National Health Service (Servicio Nacional de Salud)
SPSS: IBM SPSS Statistics

## Notes

### Competing Interest Statement

The authors have declared no competing interest.

### Author Declarations

The Florida Atlantic University Institutional Review Board reviewed and approved the protocol for this study (protocol IRB2606182).The Dominican Republic's Ministry of Public Health and Social Assistance (MISPAS) provided the data used in this analysis in anonymized, aggregate form, and the research team had no access to names, identification documents, or any other information that could directly identify patients.

